# Clinical outcomes after extended 12-month antiretroviral therapy prescriptions in a community-based differentiated HIV service delivery programme in South Africa: a retrospective cohort study

**DOI:** 10.1101/2023.03.24.23287654

**Authors:** Lara Lewis, Yukteshwar Sookrajh, Johan van der Molen, Thokozani Khubone, Phelelani Sosibo, Munthra Maraj, Rose van Heerden, Francesca Little, Reshma Kassanjee, Nigel Garrett, Jienchi Dorward

## Abstract

**Introduction:** There is an urgent need for more efficient models of differentiated antiretroviral therapy (ART) delivery for people living with HIV (PLHIV), with the World Health Organization calling for evidence to guide whether annual ART prescriptions and consultations (12M scripts) should be recommended in global guidelines. We assessed the association between 12M scripts (allowed temporarily during the COVID-19 pandemic) versus standard 6-month prescriptions and clinical review (6M scripts) and clinical outcomes.

**Methods:** We performed a retrospective cohort study using routine, de-identified data from 59 public clinics in KwaZulu-Natal, South Africa. We included PLHIV aged ≥18 years with a recent suppressed viral load (VL) who had been referred for community ART delivery with 6M or 12M scripts. We used modified Poisson regression to compare 12-month retention-in-care (not >90 days late for any visit) and viral suppression (<50 copies/mL) between prescription groups.

**Results:** Among 27,148 PLHIV referred for community ART between Jun-Dec 2020, 42.6% received 6M scripts and 57.4% 12M scripts. The median age was 39 years (interquartile range [IQR] 33-46) and 69.4% were women. Age, gender, prior community ART use and time on ART were similar in the two groups. However, more of the 12M script group had a dolutegravir-based regimen (60.0% versus 46.3%). The median (IQR) number of clinic visits in the 12 months of follow-up was 1(1-1) in the 12M group and 2(2-3) in the 6M group. Retention at 12 months was 94.6% (95% confidence interval [CI] 94.2%-94.9%) among those receiving 12M scripts and 91.8% (95% CI 91.3%-92.3%) among those with 6M scripts. 17.1% and 16.9% of clients in the 12M and 6M groups were missing follow-up VL data, respectively. Among those with VLs, 91.0% (95% CI 90.5%-91.5%) in the 12M group and 89.7% (95% CI 89.0%-90.3%) in the 6M group were suppressed. After adjusting for age, gender, ART regimen, time on ART, prior community ART use and calendar month, retention (adjusted risk ratio [aRR]: 1.03, 95% CI 1.01-1.05) and suppression (aRR: 1.01, 95% CI 1.00-1.02) were similar in the prescription groups.

**Conclusions:** Wider use of 12M scripts could reduce clinic visits without impacting short-term clinical outcomes.

## Introduction

Differentiated service delivery (DSD) models for antiretroviral therapy (ART) for people living with HIV (PLHIV) have been readily adopted worldwide. These programmes aim to provide a more client-centred approach to ART delivery and increase the accessibility of ART among PLHIV who are established on treatment, while allowing additional clinical resources to be directed towards acute and unstable patients [1]. Evidence has suggested that DSD models for ART are perceived favourably by clients [2] and do not impact negatively on their clinical outcomes [3]. As such, global HIV programmes have sought to widen the eligibility and increase the efficiencies of these programmes to facilitate the continued scale-up of ART delivery to all PLHIV [4]. Recently, interest in whether reducing clinic visit frequency within DSD programmes is safe and more efficient has developed. The World Health Organization (WHO), which currently recommends 6-monthly clinical visit frequency for PLHIV established on ART, has called for evidence on the impact of less frequent clinical visits on clinical outcomes [4].

DSD models can be broadly described within four categories, namely group models managed by healthcare workers, group models managed by clients, facility-based individual models and out-of-facility or community-based individual models [5]. In South Africa, the country with the largest ART programme globally, community-based ART delivery for individuals at external pick-up points has rapidly expanded through the Centralized Chronic Medicines Dispensing and Distribution (CCMDD) programme [6, 7]. By September 2022, over 1 million PLHIV were estimated to have accessed community-based ART through this programme. In May 2020, following the onset of the COVID-19 pandemic, ART prescriptions were extended and clinic visit frequencies within the programme were reduced from 6 to 12 months, with the purpose of minimising the number of patients visiting healthcare facilities and supporting continued ART delivery during national lockdowns [8]. However, in September 2021, these emergency provisions were not renewed and therefore lapsed.

Extending ART prescriptions from 6 to 12 months within DSD programmes could provide a more convenient service for clients and potentially reduce clinic cost and workload.

However, little is known about the impact of extended ART prescriptions on clinical outcomes among PLHIV who are established on treatment [9]. Therefore, in this study we use data from a large community-based DSD programme in South Africa to investigate whether clinical outcomes among PLHIV given 12-month ART prescriptions differed from those given 6-month prescriptions.

## Methods

### Study design and setting

We performed a retrospective cohort analysis using de-identified routinely collected electronic data from 59 public clinics in the eThekwini municipality of KwaZulu-Natal, South Africa. KwaZulu-Natal is the province with the highest HIV prevalence in South Africa, with an estimated HIV prevalence of 27% amongst adults aged 15-49 years [10]. ART is provided at all public sector clinics in accordance with South African National Guidelines [11]. Viral load testing is done at 6 and 12 months after ART initiation, and annually thereafter [12].

Clients who are stable in care (i.e., on treatment for at least 6 months, virally suppressed and without tuberculosis [TB], pregnancy, or an uncontrolled chronic condition) are also able to access ART through a variety of DSD programmes through CCMDD. These include fast-tracked appointments in clinics, adherence clubs, and community-based ART delivery at external pick-up points (defined hereafter as community-based ART). In community-based ART, clients can collect ART from pick-up points such as private pharmacies and community centres in between clinic visits. Prior to May 2020, PLHIV referred into the community ART programmes were given a 6-month ART prescription, and were required to return to the clinic every 6 months for review [13]. In May 2020, to facilitate ART delivery during the COVID-19 pandemic, an amendment was made making clients in community ART programmes eligible for a 12-month prescription and review [8]. Although prescription length during this time was either 6 or 12 months in length, PLHIV collected ART every two or three months at external pick-up points, before returning to the clinic after 6 or 12 months for a new script.

### Participants

Adults on first or second-line ART referred to community-based ART with a 6-or 12-month prescription during the period from 1 June 2020 to 1 December 2020 were included in the cohort. We used the date on which the first 6-or 12-month prescription was given in the period as baseline. Clients who were identified as having TB or being pregnant at baseline, who were missing a viral load prior to baseline or whose most recent viral load was ≥50 copies per mL, were excluded from the analysis as they failed to meet eligibility criteria for community-based ART delivery. We also excluded those who had a viral load measured more than 3 months before baseline to ensure that timing of the follow-up annual viral load of clients in the sample were better aligned. Clients were followed for 15 months after baseline with the follow-up of the last cohort of clients ending in February 2022.

### Data sources and data management

The data for this study was extracted from TIER.net, an electronic register which includes demographic, clinical and clinic visit data for all clients receiving ART in the South African public sector [14]. It includes data on visits to public health clinics, referral to external pickup points as part of the community ART delivery programme within CCMDD, ART regimens, prescription lengths, and viral load measurements. Data on ART collection at external pick-up points are not included. We analysed de-identified data using R 4.0 (R Foundation for Statistical Computing, Vienna, Austria) and SAS, version 9.4 (SAS Institute Inc).

### Variables

The primary outcomes were retention-in-care and viral suppression at 12 months after baseline. A client was defined as being retained in care at 12 months if they were not more than 90 days late for any clinic visit scheduled to occur between baseline and 365 days after baseline. Clients who were transferred to another clinic or died within 12 months of baseline were assumed to be lost-to-care. Viral load outcomes were assessed among those who were retained during follow-up. A client was defined as virally suppressed if their viral load measured closest to 12 months after baseline was less than 50 copies/mL. Those with no recorded viral load measurement during follow-up were assigned a missing value for the viral suppression outcome. We also described the number of clinic visits within the follow-up period by prescription group.

The primary exposure of interest was ART prescription length. Clients were assigned to one of two exposure groups; the first included all clients assigned a 6-month prescription at baseline and the second group included all clients who received a 12-month ART prescription at baseline. Baseline variables measured in TIER.Net and considered as potential confounders to the association between prescription length and outcomes included age, gender, ART regimen, previous exposure to community ART, time on ART and calendar month.

### Statistical analysis

Cohort baseline characteristics were summarized using median and interquartile range (IQR) values for continuous variables and using frequencies and percentages for categorical ones. We used modified Poisson regression with generalized estimating equations assuming an exchangeable working correlation structure to estimate the relative risk of retention-in-care and viral suppression while accounting for clinic clustering [15]. Multivariable regression adjusted for measured confounders. We ran a post-hoc analysis that included an interaction term between exposure group and ART regimen in multivariable models. We conducted a sensitivity analysis of the retention outcome in which clients who had been transferred to another clinic within 12 months of baseline were classified as being retained in care. We also ran a sensitivity analysis for the viral load outcome which excluded clients in the 6-month prescription group who were not given another 6-month prescription at their bi-annual visit (for example, if they received a 12-month prescription at this point they were excluded from the sensitivity analysis sample).

### Ethical approval

This work was approved by University of Kwazulu-Natal Biomedical Research Ethics Committee (BE646/17), the KwaZulu-Natal Department of Health’s Provincial Health Research Ethics Committee (KZ_201807_021), the TB/HIV Information Systems (THIS) Data Request Committee, and the eThekwini Municipality Health Unit, with a waiver for informed consent for analysis of anonymised, routinely collected data.

## Results

### Cohort characteristics

Between 1 June and 1 December 2020, 62,927 PLHIV aged ≥18 years were eligible and provided with prescriptions for community-based ART programmes from 59 clinics in eThekwini (Figure 1). To ensure that timings of the annual viral load measurements were similar in the two exposure groups, we excluded 35,779 (56.8%) clients whose most recent viral load was measured more than 3 months before baseline. Overall, 27,148 clients were included in the analysis, with 57.4% receiving 12-month prescriptions and the remaining 42.6% receiving 6-month prescriptions at baseline.

**Figure 1:**
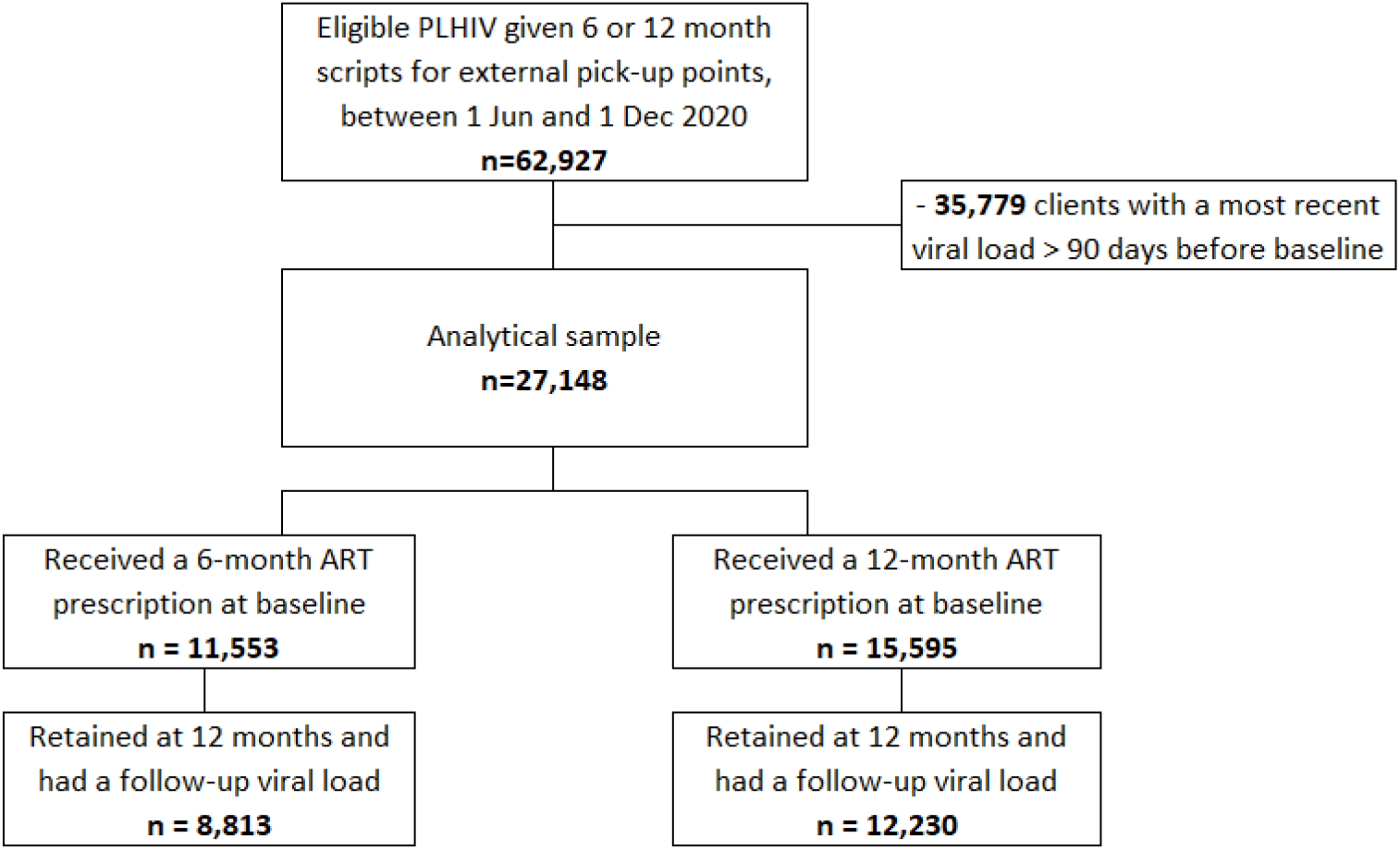
Participant flowchart.

The median age of the total cohort was 39 years (IQR 33-46) and 69.4% were women (Table 1). At baseline, the median number of years on ART of the total cohort was 5 (IQR 3-8), 97.6% were on first-line regimens and 78.5% had previously used community ART delivery. The baseline distributions of age, gender, prior community ART use and time on ART were similar in the two exposure groups. However, a larger proportion of the 12-month prescription group were on dolutegravir-based regimen (60.0%) compared to the 6-month prescription group (46.3%). The proportion of 12-month prescriptions issued varied by month (Figure 2A), with fewer being issued in November than any other month. All clinics issued a combination of 6- and 12-month prescriptions to their clients, with the median percentage of 12-month scripts issued being 61.9% (interquartile range [IQR] 44.4%-73.9%) across clinics (Figure 2B).

**Table 1:** Baseline characteristics of PLHIV referred to community ART delivery in eThekwini between June and December 2020, split by referral ART prescription length

|  |  | **Overall**<br>**(N=27,148)** | **6-month script**<br>**(N=11,553)** | **12-month script**<br>**(N=15,595)** |
| --- | --- | --- | --- | --- |
| Gender, %(n) | Male | 30.4(8242) | 29.4(3395) | 31.1(4847) |
|  | Female | 69.6(18906) | 70.6(8158) | 68.9(10748) |
| Age in years, median (IQR) |  | 39(33-46) | 39(33-46) | 39(33-46) |
| Years on ART, median (IQR) |  | 5(3-8) | 5(3-8) | 5(3-7) |
| ART regimen, %(n) | First line TLD | 54.2(14708) | 46.3(5354) | 60(9354) |
|  | First line TEE | 41.7(11331) | 48.9(5648) | 36.4(5683) |
|  | First line other | 1.7(470) | 2.1(242) | 1.5(228) |
|  | Second line | 2.4(639) | 2.7(309) | 2.1(330) |
| Previous community ART use, %(n) | No | 21.5(5840) | 21.6(2494) | 21.5(3346) |
|  | Yes | 78.5(21308) | 78.4(9059) | 78.5(12249) |
| Months since first referred to community ART, median (IQR) |  | 22(6-36) | 22(6-35) | 23(6-36) |
| Days between baseline and previous viral load, median (IQR) |  | 28(9-56) | 28(0-56) | 28(14-55) |

TLD = tenofovir disoproxil/ lamivudine/ dolutegravir; TEE = tenofovir disoproxil/ emtricitabine/ efavirenz

**Figure 2:**
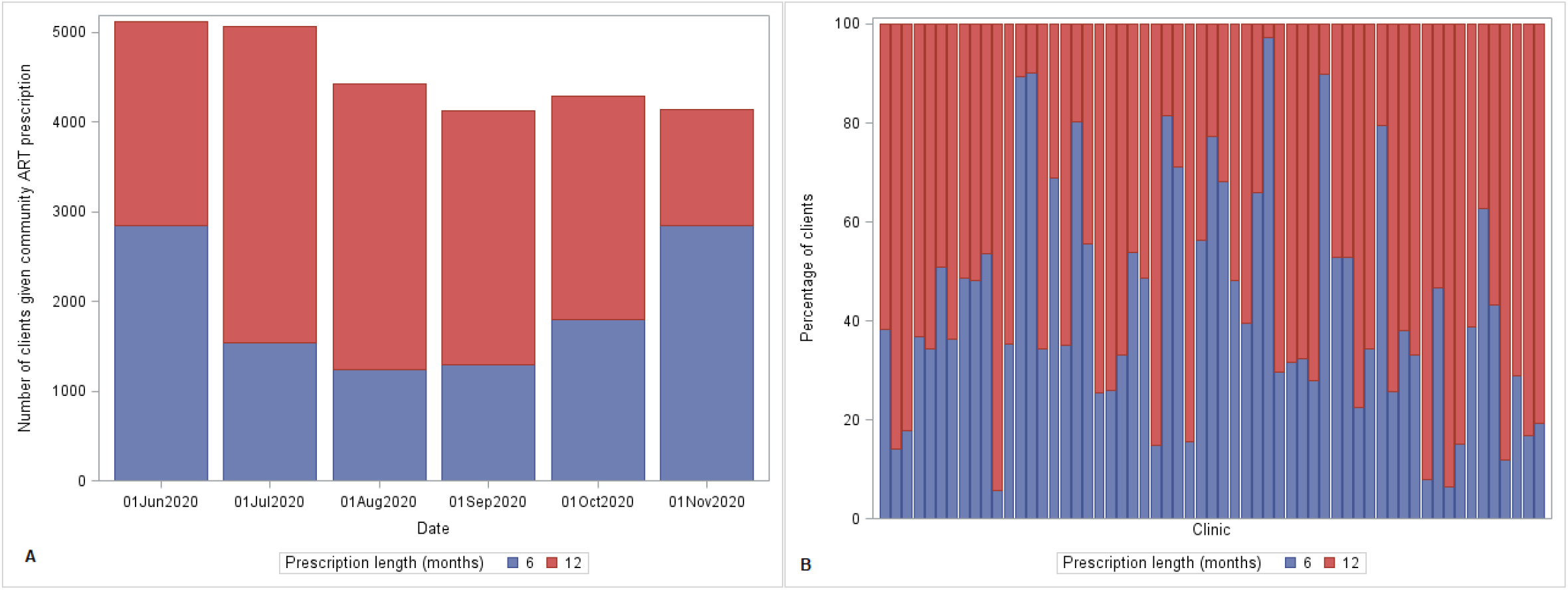
Distribution of exposure prescription groups by baseline month (A) and referring clinic (B)

### Follow-up visits

The median number of clinic visits during follow-up was 2 (IQR: 2-3) in the 6-month prescription group and 1 (IQR: 1-1) in the 12-month group. Between 6 and 12 months after baseline, 3,267 (28.3%) clients in the 6-month prescription group received a 12-month prescription and 4596 (39.8%) returned, at least temporarily, to clinic-based care.

Approximately 51.6% of those provided with 6-month prescriptions who were on a first-line regimen other than tenofovir disoproxil, lamivudine and dolutegravir (TLD) at baseline, were switched to a TLD regimen during follow-up.

### Retention in care

Of the 27,148 clients in the cohort, 1,171 (4.3%) missed a visit, 53 (0.2%) died and 565 (2.1%) were transferred to another clinic within 12 months of baseline resulting in a total retention at 12 months of 93.4% (95% confidence interval (CI) 93.1%-93.7%). Among clients receiving a 12-month and 6-month prescription at baseline respectively, 3.6% versus 5.3% missed a visit, 0.15% versus 0.25% died, and 1.6% versus 2.7% were transferred to another clinic. Overall retention was 94.6% (95% CI 94.2%-94.9%) in the 12-month prescription group and 91.8% (95% CI 91.3%-92.3%) in the 6-month group, resulting in a crude risk ratio (RR) of 1.03 (95% CI 1.02-1.04), (Table 2). After adjusting for age, gender, ART regimen, time on ART, history of community ART exposure and calendar month, the probability of retention in the 12-month prescription group was 1.03 (95% CI 1.01-1.05) times higher than in the 6-month prescription group. Retention was also positively associated with age and years on ART. Given that several clients in the 6-month prescription group changed to a TLD during follow-up, we hypothesized that the association between prescription length and retention may differ by ART regimen. However, in the post-hoc analysis including an interaction term between exposure group and ART regimen, we found no evidence of an interaction between prescription length and ART regimen (p=0.562). The association between prescription length and retention was similar in a sensitivity analysis which defined transfer-outs as retained (adjusted risk ratio [aRR]: 1.02 [95% CI 1.00-1.03], p=0.031).

**Table 2:** Retention-in-care and association with baseline characteristics of people living with HIV referred for community ART delivery in eThekwini between June and December 2020 (N=27,148)

|  |  | >90 days late for visit, %(n) | Transferred, %(n) | Died, %(n) | Retained in care, %(n) | Relative risk of retention (95% CI) | Adjusted relative risk of retention (95% CI) |
| --- | --- | --- | --- | --- | --- | --- | --- |
| Gender, %(n) | Male | 5(415) | 1.3(104) | 0.3(22) | 93.4(7701) | 1 | 1 |
|  | Female | 4(756) | 2.4(461) | 0.2(31) | 93.4(17658) | 1(0.99-1.01) | 1(1-1.01) |
| Age in years | 18-29 | 6.1(201) | 3(100) | 0.1(3) | 90.7(2977) | 1 | 1 |
|  | 30-39 | 4.7(505) | 2.4(257) | 0(2) | 92.8(9906) | 1.02(1.01-1.04) | 1.02(1.00-1.03) |
|  | 40-49 | 3.6(322) | 1.3(115) | 0.2(22) | 94.8(8430) | 1.05(1.03-1.06) | 1.04(1.02-1.05) |
|  | 50-59 | 3.3(117) | 2.1(74) | 0.5(16) | 94.1(3288) | 1.04(1.02-1.05) | 1.03(1.01-1.04) |
|  | 60+ | 3.2(26) | 2.3(19) | 1.2(10) | 93.2(758) | 1.03(1.01-1.05) | 1.02(1.00-1.04) |
| Years on ART, %(n) | <1 | 10.3(24) | 3.8(9) | 0.9(2) | 85(199) | 1 | 1 |
|  | 1-<2 | 6.1(125) | 2.9(59) | 0.1(3) | 90.9(1878) | 1.07(1.00-1.13) | 1.06(1.00-1.13) |
|  | 2-<5 | 4.9(465) | 2.1(200) | 0.2(18) | 92.8(8770) | 1.08(1.02-1.15) | 1.07(1.01-1.13) |
|  | 5+ | 3.6(557) | 1.9(297) | 0.2(30) | 94.3(14512) | 1.1(1.04-1.17) | 1.08(1.01-1.14) |
| ART regimen, %(n) | First line TLD | 4.1(599) | 1.7(255) | 0.2(31) | 94(13823) | 1 | 1 |
|  | First line TEE | 4.7(527) | 2.5(280) | 0.2(21) | 92.7(10503) | 0.99(0.98-0.99) | 0.99(0.98-1.00) |
|  | First line other | 4.7(22) | 3.4(16) | 0(0) | 91.9(432) | 0.98(0.96-1.00) | 0.98(0.96-1.00) |
|  | Second line | 3.6(23) | 2.2(14) | 0.2(1) | 94.1(601) | 1.00(0.98-1.03) | 1.00(0.98-1.03) |
| Previous community ART use, %(n) | No | 5.9(346) | 2.4(143) | 0.2(12) | 91.4(5339) | 1 | 1 |
|  | Yes | 3.9(825) | 2.0(422) | 0.2(41) | 94(20020) | 1.03(1.01-1.04) | 1.02(1.00-1.03) |

|  |  | >90 days late<br>for visit, %(n) | Transferred,<br>%(n) | Died, %(n) | Retained in<br>care, %(n) | Relative risk of<br>retention (95%<br>CI) | Adjusted<br>relative risk of<br>retention (95%<br>CI) |
| --- | --- | --- | --- | --- | --- | --- | --- |
| Calendar month | June 2020 | 3.7(189) | 2.1(107) | 0.2(10) | 94(4812) | 1 | 1 |
|  | July 2020 | 3.9(200) | 2(101) | 0.2(12) | 93.8(4757) | 1.00(0.99-1.01) | 0.99(0.98-1.00) |
|  | August 2020 | 3.8(167) | 2(88) | 0.2(10) | 94(4153) | 1.00(0.99-1.01) | 0.99(0.98-1.00) |
|  | September 2020 | 4.7(193) | 2.1(86) | 0.2(8) | 93(3836) | 0.99(0.98-1.00) | 0.98(0.97-1.00) |
|  | October 2020 | 4.9(210) | 2.2(95) | 0.1(6) | 92.7(3972) | 0.99(0.97-1.00) | 0.98(0.97-1.00) |
|  | November 2020 | 5.1(212) | 2.1(88) | 0.2(7) | 92.6(3829) | 0.99(0.97-1.00) | 0.99(0.97-1.01) |
| ART prescription length | 6 months | 5.2(606) | 2.7(309) | 0.3(29) | 91.8(10609) | 1 | 1 |
|  | 12 months | 3.6(565) | 1.6(256) | 0.2(24) | 94.6(14750) | 1.03(1.02-1.04) | 1.03(1.01-1.05) |

TLD = tenofovir disoproxil/ lamivudine/ dolutegravir; TEE = tenofovir disoproxil/ emtricitabine/ efavirenz

### Viral load outcomes

Among clients receiving a 6-month and 12-month prescription at baseline respectively, 83.1% and 82.9% had a follow-up viral load result recorded at a median of 12 (IQR 10-12) and 12 (IQR 11-12) months after baseline (Table 3). At follow-up, viral suppression was 91.0% (95% CI 90.5%-91.5%) in the 12-month prescription group compared to 89.7% (95% CI 89.0%-90.3%) in the 6-month prescription group (RR 1.01, 95% CI 1.00-1.02, p=0.008). After adjusting for age, gender, time on ART, ART regimen, history of community ART exposure and calendar month, viral suppression was not found to be significantly associated with whether a 6-or 12-month prescription had been given (aRR 1.01, 95% CI 1.00-1.02, p=0.263). In the post-hoc analysis including an interaction term between prescription length and ART regimen, the interaction term was not significantly associated with viral suppression suggesting that ART regimen did not alter the association between prescription group and viral suppression (p=0.803). In the sensitivity analysis in which clients who were given prescriptions at their bi-annual visit of any length other than 6 months were removed, results were similar to the primary analysis, as the likelihood of viral suppression in the 12-month prescription group was not significantly different from that in the 6-month group (aRR 1.01, 95% CI 1.00-1.03, p=0.073).

**Table 3:** Viral suppression and association with baseline characteristics of people living with HIV referred for community ART delivery in eThekwini between June and December 2020 (N= 25,359)

|  |  | Missing viral load, %(n) | Virally suppressed*, %(n) | Relative risk of suppression* (95% CI) | Adjusted relative risk of suppression* (95% CI) |
| --- | --- | --- | --- | --- | --- |
| Gender, %(n) | Male | 17.4(1339) | 88.4(5624) | 1 | 1 |
|  | Female | 16.9(2977) | 91.3(13408) | 1.04(1.02-1.05) | 1.03(1.02-1.04) |
| Age in years | 18-29 | 17.3(516) | 91.1(2242) | 1 | 1 |
|  | 30-39 | 17.2(1701) | 90.8(7450) | 0.99(0.98-1.01) | 1.00 (0.99-1.01) |
|  | 40-49 | 16.8(1414) | 90.3(6333) | 0.99(0.97-1.00) | 1.00 (0.98-1.01) |
|  | 50-59 | 16.6(546) | 89.4(2450) | 0.98(0.96-0.99) | 0.99(0.97-1.00) |
|  | 60+ | 18.3(139) | 90(557) | 0.99(0.96-1.02) | 1.00 (0.97-1.03) |
| Years on ART, %(n) | <1 | 27.1(54) | 83.4(121) | 1 | 1 |
|  | 1-<2 | 16.7(313) | 91.9(1438) | 1.10(1.03-1.17) | 1.09(1.02-1.17) |
|  | 2-<5 | 17.6(1546) | 90.5(6538) | 1.08(1.01-1.16) | 1.07(0.99-1.14) |
|  | 5+ | 16.6(2403) | 90.3(10935) | 1.08(1.01-1.15) | 1.06(0.99-1.14) |
| ART regimen, %(n) | First line TLD | 17.1(2366) | 90.5(10365) | 1 | 1 |
|  | First line TEE | 16.8(1769) | 91(7945) | 1.01(1.01-1.02) | 1.01(1.00-1.02) |
|  | First line other | 14.1(61) | 90.8(337) | 1.01(0.98-1.04) | 1.01(0.98-1.05) |
|  | Second line | 20(120) | 80(385) | 0.89(0.85-0.93) | 0.89(0.85-0.93) |
| Previous community ART use, %(n) | No | 16.6(886) | 89.7(3994) | 1 | 1 |
|  | Yes | 17.1(3430) | 90.6(15038) | 1.01(1.00-1.02) | 1.02(1.00-1.03) |
| Calendar month | June 2020 | 15.1(725) | 90.8(3712) | 1 | 1 |
|  | July 2020 | 16.4(782) | 90.7(3605) | 1.00(0.99-1.02) | 1.00(0.99-1.01) |
|  | August 2020 | 16.2(673) | 92.2(3210) | 1.02(1.00-1.03) | 1.01(1.00-1.03) |
|  | September 2020 | 16.2(623) | 92(2955) | 1.01(1.00-1.03) | 1.01(0.99-1.03) |
|  | October 2020 | 18.2(723) | 89.2(2897) | 0.98(0.96-1.00) | 0.98(0.96-1.00) |
|  | November 2020 | 20.6(790) | 87.3(2653) | 0.96(0.94-0.98) | 0.96(0.94-0.98) |
| ART prescription length | 6 months | 16.9(1796) | 89.7(7903) | 1 | 1 |
|  | 12 months | 17.1(2520) | 91(11129) | 1.01(1.00-1.02) | 1.01(1.00-1.02) |

TLD = tenofovir disoproxil/ lamivudine/ dolutegravir; TEE = tenofovir disoproxil/ emtricitabine/ efavirenz; \*excluding missing viral loads from calculation (N=21,043)

## Discussion

Using a large cohort of PLHIV referred for community-based ART delivery, we found that clinical outcomes among PLHIV receiving 12-month ART prescriptions were similar to those among PLHIV receiving 6-month prescriptions. In addition, those provided with 12-month prescriptions had half the number of clinic visits during the 12 months of follow-up compared to those with 6-month prescriptions.

Our findings are consistent with those from existing research, although data are lacking. A recent systematic review compared clinical outcomes among PLHIV with reduced (6-to 12-monthly) clinical consultations to those among PLHIV with 3-monthly clinical visits [9]. All studies included in the review analysed PLHIV who were established on ART, and most utilised clinical outcomes with a 12-month duration. The authors showed no difference in retention among clients on 6- and 12-monthly clinic visit schedules compared to those on 3-monthly ones, and these findings were consistent across delivery strategies. However, the results for viral suppression outcomes were, overall, inconclusive. Moreover, few studies compared outcomes among those with 12-monthly clinical visits directly to those with 6-monthly visits.

Although clinical outcomes may be similar, moving from 6-monthly to annual prescriptions may be beneficial to clients and providers in other ways. Qualitative and preference data has indicated that PLHIV on treatment have a strong preference for longer intervals between clinic visits [16-21]. Despite the value of psychosocial support obtained through face-to-face clinical interaction, the benefits of reduced travel time, opportunity costs and incidents of unintended disclosure achieved through less frequent clinical visits are perceived as considerable among clients. There is however little data on client’s perceived benefits of moving beyond 6-monthly visits, although one study has suggested that clients prefer 6-monthly cycles to less and more frequent ones [20]. For providers, some studies have shown that reduced visit frequency can reduce clinic congestion and workload and allow more time for unstable or acute patients [22, 23]. This is supported by the results in our study that showed the number of clinic visits was halved when clients moved from 6-monthly to annual consultations.

Although our analysis controlled for several potential confounders, it is possible that there were unmeasured confounders which, through their exclusion, would have biased our results. Of key concern was whether people who were perceived by clinicians to be more clinically stable would have been more likely to be given a 12-month prescription. Within the parameters available in the dataset, all clients regardless of prescription group had a recent suppressed viral load, did not have TB and were not pregnant at baseline, and as such had been referred for community-based ART. We were unable to investigate why some clients were prioritised for annual prescriptions over others if not for differences in clinical stability. Potential reasons could be that some clients preferred more frequent visits or that healthcare workers selected prescription length to ensure clients’ clinic schedules did not clash with the national holidays in December. The period of observation occurred during the national roll-out of dolutegravir [24], and this may have impacted on clinic staff’s willingness to provide long prescriptions for people on efavirenz, as they may have anticipated needing to transition to dolutegravir in the future. We noted that those receiving 12-month prescriptions were more likely to be on a dolutegravir-regimen and that more than half of the clients starting on TEE provided with 6-month prescriptions had moved to TLD by the end of follow-up.

The period of observation used in this study occurred at the beginning of the COVID-19 pandemic in South Africa, meaning our results may not be generalisable to non-pandemic situations. However, we found no difference in outcomes by baseline time period, even though the SARS-CoV-2 Beta variant wave occurred during the second half of the baseline time period. Another limitation was that, while we aimed to compare clinical outcomes of those with 12-month prescriptions to those with 6-month prescriptions, more than 20% of clients initially provided with a 6-month prescription received a 12-month prescription before the end of follow-up. We were unable to perform a “per-protocol” type analysis for the retention-in-care outcome because by removing these clients we would have introduced bias by excluding clients who we know were retained at 6 months. Lastly, another COVID-19 adaptation introduced in South Africa was the option of automatic 6-month re-scripting, meaning that a client with an existing 6-month prescription could have a new 6-month prescription prescribed remotely, without an in-person clinical consultation. While this was not common at the study clinics, it was not well recorded in TIER.net, meaning that some people in the 6-month script group may have actually had their second script issued remotely, rather than in person. If these people had worse clinical outcomes, it could bias the estimates in the 6 month group to be worse. However, we still demonstrate that outcomes in the 12-month group remained very good, with retention in care of 94.6% and viral suppression 91.0%.

While the findings from this study are reassuring, more needs to be understood about the long-term clinical outcomes of extending ART prescriptions from 6 to 12 months and whether these findings translate to different populations such as children and adolescents, and those outside of South Africa. Moreover, future research should examine whether good clinical outcomes in those with extended 12-month prescriptions are retained if multi-month dispensing increases beyond 2 or 3 months (the frequency used in this analysis). Qualitative research examining clients’ perceptions of moving from 6-to 12-monthly ART prescriptions also needs to be better understood, as well as provider’s perceptions. Finally, the impact of on clinic workload needs to be better quantified.

## Conclusions

In this study, we demonstrate that extending ART prescriptions in community-based ART delivery programmes from 6 to 12 months is associated with better retention-in-care and similar viral suppression after 12 months. If annual prescriptions do not negatively impact on cost or clinic workload and are perceived favourably by clients, they should be considered for DSD programmes to facilitate the rapid scale-up of DSD worldwide.

## Data Availability

The data used for this analysis cannot be shared publicly because of legal and ethical requirements regarding use of routinely collected clinical data in South Africa. Researchers may request access to the data from the eThekwini Municipality Health Unit (contact details obtainable upon request to corresponding author).

## Authorship contributions

JD, NG and LL conceived the analysis. TK, PS, YS, MM and RvH oversaw data collection. TK and JvdM oversaw data curation. TK, JvdM, LL and JD have verified the underlying data. LL, JvdM and JD analysed the data with inputs on design and implementation from RK and FL. LL drafted the manuscript. All authors critically reviewed and edited the manuscript and consented to final publication.

## Acknowledgements

We would like to thank the staff and patients at eThekwini Municipality Health Unit primary care clinics, as well as Riona Govender and Sifiso Pakhati from the Health Informatics Directorate in the National Department of Health.

## References

1. Grimsrud A, Bygrave H, Doherty M, Ehrenkranz P, Ellman T, Ferris R, et al. Reimagining HIV service delivery: the role of differentiated care from prevention to suppression. J Int AIDS Soc. 2016;19(1):21484.

2. Dorward J, Msimango L, Gibbs A, Shozi H, Tonkin-Crine S, Hayward G, et al. Understanding how community antiretroviral delivery influences engagement in HIV care: a qualitative assessment of the Centralised Chronic Medication Dispensing and Distribution programme in South Africa. BMJ Open. 2020;10(5):e035412.

3. Long L, Kuchukhidze S, Pascoe S, Nichols BE, Fox MP, Cele R, et al. Retention in care and viral suppression in differentiated service delivery models for HIV treatment delivery in sub-Saharan Africa: a rapid systematic review. J Int AIDS Soc. 2020;23(11):e25640.

4. Consolidated Guidelines on HIV Prevention, Testing, Treatment, Service Delivery and Monitoring: Recommendations for a Public Health Approach. WHO Guidelines Approved by the Guidelines Review Committee. Geneva 2021.

5. Grimsrud A, Barnabas RV, Ehrenkranz P, Ford N. Evidence for scale up: the differentiated care research agenda. J Int AIDS Soc. 2017;20(Suppl 4):22024.

6. Liu L, Christie S, Munsamy M, Roberts P, Pillay M, Shenoi SV, et al. Expansion of a national differentiated service delivery model to support people living with HIV and other chronic conditions in South Africa: a descriptive analysis. BMC health services research. 2021;21(1):1–8.

7. Health Systems Trust. CCMDD [17 October 2022]. Available from: https://getcheckedgocollect.org.za/ccmdd/.

8. South African National Department of Health. Government Gazette No. 43294:514. 2020.

9. Le Tourneau N, Germann A, Thompson RR, Ford N, Schwartz S, Beres L, et al. Evaluation of HIV treatment outcomes with reduced frequency of clinical encounters and antiretroviral treatment refills: A systematic review and meta-analysis. PLoS Med. 2022;19(3):e1003959.

10. Human Sciences Research Council. South African National HIV Prevalence, Incidence, Behaviour and Communication Survey, 2017 Cape Town, South Africa: Human Sciences Research Council,; 2018.

11. South African National Department of Health. Adherence Guidelines for HIV, TB and NCDS: Standard Operating Procedures - 2020. National Department of Health; 2020.

12. South African National Department of H. National consolidated guidelines for the prevention of mother to child transmission of HIV (PMTCT) and the management of HIV in children, adolescents and adults. Pretoria, South Africa; 2015.

13. Meyer JC, Schellack N, Stokes J, Lancaster R, Zeeman H, Defty D, et al. Ongoing initiatives to improve the quality and efficiency of medicine use within the public healthcare system in South Africa; A preliminary study. Frontiers in Pharmacology. 2017;8(NOV):1–16.

14. Osler M, Hilderbrand K, Hennessey C, Arendse J, Goemaere E, Ford N, et al. A three-tier framework for monitoring antiretroviral therapy in high HIV burden settings. Journal of the International AIDS Society. 2014;17(1):18908-.

15. Yelland LN, Salter AB, Ryan P. Performance of the modified Poisson regression approach for estimating relative risks from clustered prospective data. Am J Epidemiol. 2011;174(8):984–92.

16. Zanolini A, Sikombe K, Sikazwe I, Eshun-Wilson I, Somwe P, Bolton Moore C, et al. Understanding preferences for HIV care and treatment in Zambia: Evidence from a discrete choice experiment among patients who have been lost to follow-up. PLoS Med. 2018;15(8):e1002636.

17. Keene CM, Zokufa N, Venables EC, Wilkinson L, Hoffman R, Cassidy T, et al. ‘Only twice a year’: a qualitative exploration of 6-month antiretroviral treatment refills in adherence clubs for people living with HIV in Khayelitsha, South Africa. BMJ Open. 2020;10(7):e037545.

18. Eshun-Wilson I, Mukumbwa-Mwenechanya M, Kim HY, Zannolini A, Mwamba CP, Dowdy D, et al. Differentiated Care Preferences of Stable Patients on Antiretroviral Therapy in Zambia: A Discrete Choice Experiment. J Acquir Immune Defic Syndr. 2019;81(5):540–6.

19. Zakumumpa H, Makobu K, Ntawiha W, Maniple E. A mixed-methods evaluation of the uptake of novel differentiated ART delivery models in a national sample of health facilities in Uganda. PLoS One. 2021;16(7):e0254214.

20. Dommaraju S, Hagey J, Odeny TA, Okaka S, Kadima J, Bukusi EA, et al. Preferences of people living with HIV for differentiated care models in Kenya: A discrete choice experiment. PLoS One. 2021;16(8):e0255650.

21. Strauss M, George G, Mantell JE, Mapingure M, Masvawure TB, Lamb MR, et al. Optimizing Differentiated HIV Treatment Models in Urban Zimbabwe: Assessing Patient Preferences Using a Discrete Choice Experiment. AIDS Behav. 2021;25(2):397–413.

22. Hubbard J, Phiri K, Moucheraud C, McBride K, Bardon A, Balakasi K, et al. A Qualitative Assessment of Provider and Client Experiences With 3-and 6-Month Dispensing Intervals of Antiretroviral Therapy in Malawi. Glob Health Sci Pract. 2020;8(1):18–27.

23. Prust ML, Banda CK, Nyirenda R, Chimbwandira F, Kalua T, Jahn A, et al. Multi-month prescriptions, fast-track refills, and community ART groups: results from a process evaluation in Malawi on using differentiated models of care to achieve national HIV treatment goals. J Int AIDS Soc. 2017;20(Suppl 4):21650.

24. South African National Department of Health. 2019 ART Clinical Guidelines for the management of HIV in Adults, Pregnancy, Adolescents, Children, Infants and Neonates.. South Africa; 2019.

